# Disease-specific contribution of pulvinar dysfunction to impaired emotion recognition in schizophrenia

**DOI:** 10.1101/2021.05.21.21257528

**Authors:** Antígona Martínez, Russell H. Tobe, Pablo A. Gaspar, Daniel Malinsky, Elisa C. Dias, Pejman Sehatpour, Peter Lakatos, Gaurav H. Patel, Gail Silipo, Daniel C. Javitt

**Affiliations:** Nathan Kline Institute for Psychiatric Research, Orangeburg, NY, USA; Columbia University, College of Physician and Surgeons, New York, NY, USA; Department of Psychiatry, Biomedical Neurosciences Institute, IMHAY, University of Chile, Santiago, Chile; Columbia University, Mailman School of Public Health, New York, NY, USA; New York State Psychiatric Institute, New York, NY, USA

## Abstract

One important aspect for managing social interactions is the ability to rapidly and accurately perceive and respond to facial expressions, which is highly dependent upon intact processing within both cortical and subcortical components of the early visual pathways. Social cognitive deficits, including face emotion recognition (FER) deficits, are characteristic of several neuropsychiatric disorders, including schizophrenia (Sz) and autism spectrum disorders (ASD). Here, we investigated potential visual sensory contributions to FER deficits in Sz (n=28) and adult ASD (n=20) participants compared to neurotypical (n=30) controls using task-based fMRI during an implicit static/dynamic FER task. Compared to neurotypical controls, both Sz and ASD participants had significantly lower FER scores which interrelated with diminished activation of the superior temporal sulcus (STS). In Sz, STS deficits were predicted by reduced activation of both early visual regions and the pulvinar nucleus of the thalamus, along with impaired cortico-pulvinar interaction. By contrast, ASD participants showed patterns of increased early visual cortical and pulvinar activation. Large effect-size structural and histological abnormalities of pulvinar have previously been documented in Sz. Moreover, we have recently demonstrated impaired pulvinar activation to simple visual stimuli in Sz. Here, we provide the first demonstration of a disease-specific contribution of impaired pulvinar activation to social cognitive impairment in Sz.

## Introduction

Social cognitive deficits are a core feature of schizophrenia (Sz)^1^ and autism spectrum disorders (ASD) and contribute to impaired functional outcome^2,3^. One important aspect of social functioning is the ability to rapidly and accurately perceive facial expressions. Impaired face-emotion recognition (FER) has been extensively reported in Sz^4,5^ and ASD^6,7^ however the underlying neuronal substrates of these deficits are not fully understood and, indeed, may arise from differential underlying neural pathologies^8^. Over recent years, the contribution of sensory-processing deficits to cognitive impairments has been increasingly appreciated, including the potential role of dysfunction within subcortical components of the afferent visual streams. Here, we utilize functional magnetic resonance imaging (fMRI) to evaluate the contributions of impaired early sensory processing to FER impairments in Sz. Data were collected as well from both neurotypical and ASD comparison groups in order to assess the specificity and magnitude of observed activation deficits in Sz.

During normative brain function, FER is supported by activation of specific components of the “social brain”^9^, which includes structures along both the dorsal and ventral visual-cortical pathways^10-12^. These pathways receive retinal information from the lateral geniculate nucleus (LGN), which projects to primary visual cortex (V1). The dorsal pathway receives its primary input from the magnocellular geniculostriate pathway and is specialized for rapid detection of low spatial-frequency and motion information. Key dorsal structures include motion-sensitive mid-temporal regions (MT, MST). In Sz, differential deficits in magnocellular processing have been reported and related to potentially impaired patterns of sensory gain and functions of the N-methyl-D-aspartate-type glutamate receptors (NMDAR) (reviewed in^13^). Moreover, impairments in magnocellular function correlate with behavioral measures of impaired FER in Sz, supporting the involvement of this pathway in social cognition^14-16^. By contrast, the ventral visual stream receives predominant input from the subcortical parvocellular system and is specialized for slower but higher-resolution processing of stimulus details. Key targets of the ventral stream include visual area V4 and the fusiform face complex (FFC).

An additional cortical area important for processing social stimuli is the superior temporal sulcus (STS), which has been reliably associated with processing biological motion signals^17-19^ including the dynamic changeable aspects of facial features (eyes, lips). Impaired STS activation has been documented in Sz but the basis for the deficit remains unknown^23-27^.

In addition to the cortical system, humans retain an evolutionarily old retinogeniculate system that mediates non-conscious affective processing via amygdala, superior colliculus and the pulvinar nucleus of the thalamus (PulN) (reviewed in^28^). In addition to mediating retinogeniculate input into visual cortex, PulN also mediates cortico-cortical interactions between successive brain regions within the dorsal and ventral stream pathways (e.g.^29^), and is the site of greatest NMDAR density within primate thalamus^30^. PulN is anatomically divided into discrete, functionally differentiated subdivisions (e.g.^31,32^). For example, the ‘visual pulvinar’, consisting of its inferior (PI) and lateral (PL) subdivisions, has dense connections with early visual sensory regions^32^ and likely plays a modulatory role in visual information processing^33^. In addition, projections from PI specifically innervate motion sensitive regions surrounding area MT, especially MST^34^, and also serve as drivers to secondary areas of visual cortex (e.g. V2) and as modulators to V1^33^. On the other hand, medial pulvinar (PM) is considered multimodal and is primarily coupled with prefrontal and temporal regions including STS^35^ and may play a unique role in processing emotional information (reviewed in^36^).

Here, we evaluate whole-brain fMRI activation patterns during FER in Sz, relative to both neurotypical individuals and ASD. Inclusion of ASD controls is based on a previous study involving simple visual stimuli in which we observed a divergent pattern of disturbance within early visual areas and PulN relative to Sz, despite similar magnitude of FER impairment^14^. We hypothesized that, in Sz, deficits within FER-related higher tier visual regions (e.g. STS) would be driven significantly by impaired activation within both early visual regions (e.g. V1, MST) and PulN as well as impaired cortico-pulvinar interaction. Moreover, we hypothesized that deficit patterns would be differential across Sz and ASD despite similar levels of behavioral impairment, suggesting disorder-specific pathophysiological mechanisms underlying social cognitive impairments across neuropsychiatric populations.

## Materials and Methods

### Participants

Seventy-eight participants took part, including 28 participants diagnosed with Sz and on stable doses of antipsychotic medication, 30 neurotypical controls and 20 adults with ASD (**Supplementary Methods**). The investigation was approved by the Nathan Kline Institute (NKI) institutional review board. Informed consent was obtained from all participants.

### Paradigm

Unique video clips of five actors (three male) dynamically expressing each of four emotions (happy, sad, angry, fearful) were selected from the University of Cambridge Mind Reading Emotions Library (adult level 6)^37^. Five additional actors (two male) from the NKI community acted a neutral expression consisting of non-emotionally salient head/eye movements (left/right, up/down). Neutral videos were matched in size, resolution and luminance to emotion videos. For each video, representative single frames were extracted and used as corresponding static stimuli. Both dynamic and static stimuli were presented for 2s each. In each of two 7.5-minute fMRI scans, dynamic and static stimuli of a single emotion/neutral were delivered in 12-second blocks interleaved with 10-seconds of fixation-only. A total of ten blocks of static/dynamic faces either emotional or neutral were presented in random order per scan (**Figure 1A**). Participants responded to a predesignated actor, irrespective of emotion or motion.

**Figure 1.**
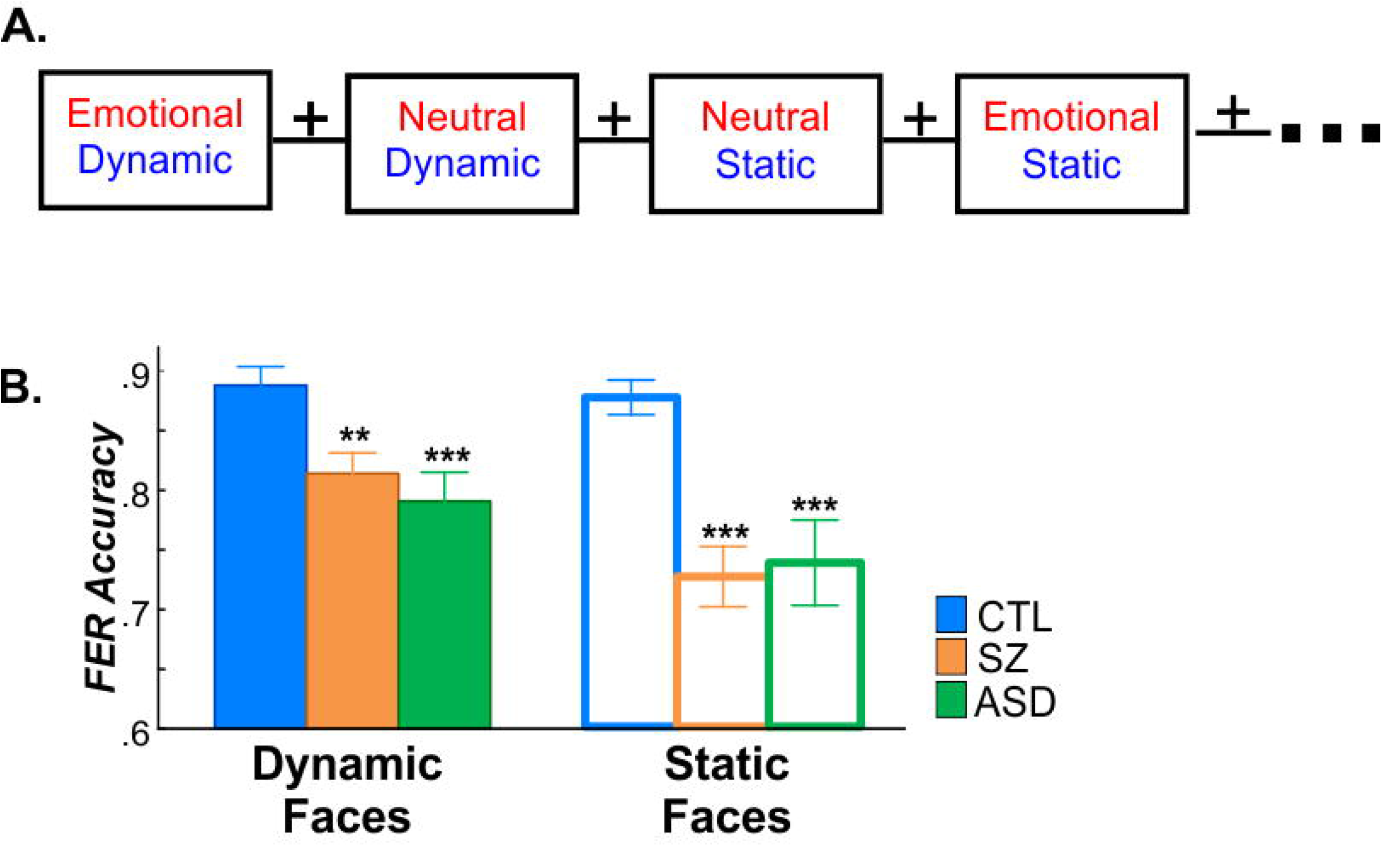
**A**. Schematic of fMRI paradigm. **B**. FER accuracy for dynamic (filled bars) and static (open) faces in control (CTL; blue), schizophrenia (SZ; orange) and autism (ASD; green). Significance of group differences are relative to CTL (*p<.05; **p<.01;***p<.005).

### Behavior measures

Following scanning, a forced-choice task was administered outside the scanner using static and dynamic emotional faces, along with the Penn Emotion Recognition (ER-40) test^38^. For both tests, percent correct responses were recorded (see **Supplementary Methods**).

### Functional Imaging

Imaging took place on a Siemens 3T TiM Trio scanner. Functional MRI data were preprocessed and analyzed with Analysis of Functional NeuroImages (AFNI) software^39,40^ (see **Supplementary Methods** for acquisition/analyses details). Individual cortical surfaces were rendered from high-resolution anatomical images using Freesurfer and registered to the fsaverage mesh^41^. The pulvinar and amygdala were derived individually using a Bayesian atlas-based automated segmentation method^42-44^. Primary analyses involved the entire pulvinar. In secondary analyses, pulvinar subdivisions were tested separately. Single-subject data were analyzed on the cortical surface with a general linear model including stimulus-specific regressors and head-motion parameters. Identical analyses were carried out in native-space volumes to assess subcortical structures. The HCP-MMP1.0 cortical parcellation^45^, resampled to fsaverage, was applied to individual data (**Supplementary Figure 1A**). For group-level analyses, mean beta parameter estimates were extracted from cortical parcels of interest and from pulvinar and amygdala.

### General statistics

Across- or between-group comparisons of beta parameter effect sizes were conducted using univariate (ANOVA) or multivariate (MANOVA) repeated measures analysis of variance as appropriate, with a between-group factor of diagnostic group and within-group factors of face-motion (dynamic, static) and face-emotion (emotional, neutral). In secondary analyses, effects were analyzed as a function of face-emotion-type (happy, sad, angry, fearful). The relationships among measures were assessed by analysis of covariance (ANCOVA) or by Pearson correlation. All statistics were two-tailed with preset α threshold for significance set to 0.05.

### Path Analyses

Exploratory linear mediation analyses were carried out using the PROCESS3.5 macro^46^ implemented in SPSS26 (https://www.ibm.com/products/spss-statistics). Statistical significance of indirect pathways, reflecting the impact of mediation, was evaluated using a non-parametric bootstrap approach with 10,000 replication samples to obtain a 95% confidence interval (CI)^47^.

## Results

### Behavior

Mean FER accuracy on the dynamic/static FER task was lower in Sz (F(1,56)=15.02, p<.001) and ASD participants (F(1,48)=7.67, p=.009) than neurotypical controls (**Figure 1B, Supplementary Figure S2A**) and intercorrelated across subjects with ER-40 test results (F(1,72)=8.01, p=.006; R^2^=.401) (**Supplementary Figure S2B**,**C**). Behavioral performance on the identity recognition task during fMRI was marginally worse in Sz (F(1,56)=3.76, p=.058) and significantly worse in ASD (F(1,48)=4.52, p=.039) compared to controls.

### Between-group activation differences (Sz vs. control)

To avoid issues related to circularity in data analysis^48^, parcels of interest were first identified across all participants, yielding a mask consisting of 35 bilateral parcels (**Supplementary Figure S1B**,**C**). Within this mask the group X parcel (F(34,23)=2.14, p=.030) interaction was significant, prompting follow-up analyses.

Compared to controls, reduced activation (beta parameter estimates) were observed in Sz patients within seven parcels comprising primary (V1, p=.002), early (V4, p=.005), motion-sensitive (MST, p=.042), face-selective (FFC, p=.045), superior-temporal (TPOJ1, p=.001; STSdp, p=.019) and inferior-frontal (IFSp, p=.041) regions (**Figure 2A-D**; **Supplementary Table 1**). Overall, activations were larger over the right hemisphere (F(1,56)=24.12, p<.001) but were equivalently so for Sz and control participants (F(1,56)=.28, p=.597). Subsequent tests were collapsed across hemispheres.

**Figure 2.**
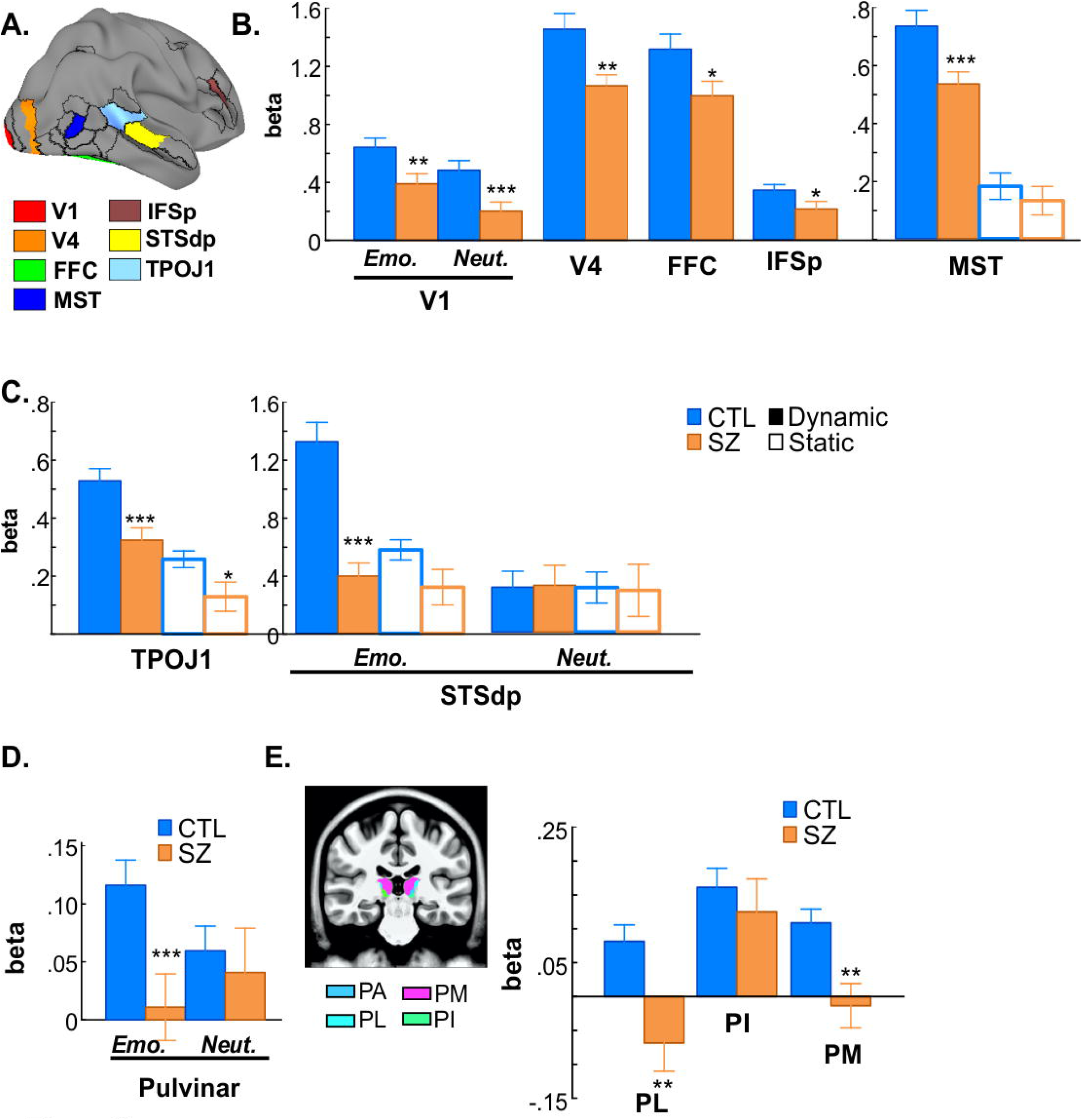
**A**. Localization of key parcels. **B**. Mean activation (beta parameter) for CTL and SZ groups. For areas with face-motion effects, activation to dynamic (solid bars)/static (open bars) faces are plotted separately. **C**. Activation in STS parcels including differential patterns in STSdp. **E**. Whole-pulvinar activation for emotional/neutral faces. **F**. Localization and groupwise activations of lateral (PL), inferior (PI) and medial (PM) pulvinar subdivisions.

A significant across-group effect of face-emotion was observed in V1 (F(1,56)=4=17.21, p<.001) and, as expected, of face-motion in MST (F(1,56)=132.92, p<.001) (**Figure 2B)**. In MST, the group X face-motion interaction was significant (F(1,56)=4.08, p=.048), reflecting reduced activation in Sz specific to dynamic (p=.005) but not static (p=.460) faces, irrespective of face-emotion (**Figure 2B)**.

A main effect of face-motion was also observed in both STS parcels (TPOJ1: (F(1,56)=4.160, p<.001); STSdp: (F(1,56)=13.82, p<.001). In STSdp, the main effects of face-emotion (F(1,56)=10.35, p=.002) was significant, as was the group X face-motion (F(1,56)=5.13, p=.027), group X face-emotion, (F(1,56)=12.31, p=.001) and group X face-motion X face-emotion (F(1,56)=5.28, p=.025) interactions, reflecting preferential activation deficits to dynamic emotional faces (t(56)=5.69, p<.001) (**Figure 2C**).

### Pulvinar

In PulN, activations were reduced in Sz overall (F(1,56)=4.35, p=.042) especially to emotional versus neutral faces (F(1,56)=4.96, p=.030) (**Figure 2D**). All other main effects and interactions were non-significant (p >.2 for all).

When PulN subdivisions were analyzed separately, mean activations in the lateral (PL) (F(1,56)=9.97, p=.003) and medial (PM) (F(1,56)=10.35, p=.002) subnuclei were significantly reduced in Sz. Activation of the inferior subdivision (PI) did not differ significantly between groups (F(1,56)=.43, p=.513) (**Figure 2E**).

### Regression and Path Analyses

The interrelationship between cortical and subcortical activation deficits in Sz was assessed by ANCOVA and mediation tests. FER scores were first covaried against mean activation of all cortical/subcortical areas **(see Supplementary Fig. S2B)**. Activation of STSdp predicted FER scores across groups (F(1,54)=17.03, p<.001; Adj. R^2^=.611) and in Sz (r=.542, p=.003) and control (r=.470, p=.009) groups independently (**Figure 3A**). In turn, activation of both V1 (F(1,54)=7.13, p=.010; Adj. R^2^=.254) and TPOJ1 (F(1,54)=9.55, p=.003; Adj. R^2^=.275) predicted STSdp activity (**Figure 3B**). Finally, in all subjects, MST activation was a strong predictor of TPOJ1 (F(1,54)=28.65, p<.001 Adj. R^2^=.472) (**Figure 3C**). In all cases, the covariate X group interaction was non-significant suggesting a similar slope of the relationships across groups.

**Figure 3.**
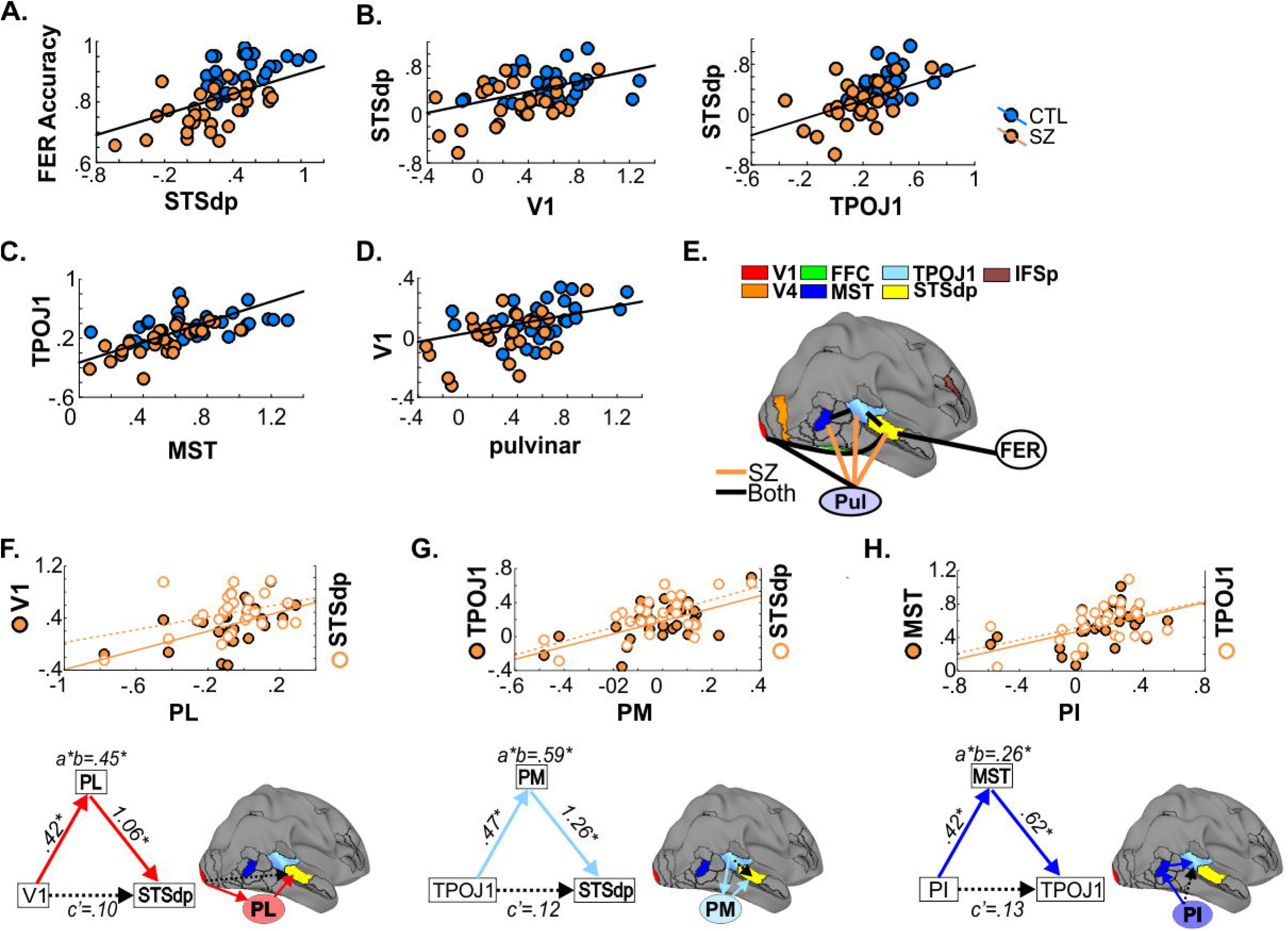
**A**. Groupwise correlation between STSdp activation and FER performance. **B**. Correlation between STSdp-V1 and STSdp-TPOJ1 activation; **C**. MST-TPOJ1 correlation; **D**. Whole-pulvinar-V1 correlation. **E**. Interrelationship between activated cortical regions and FER. Black lines: significant relationship across groups. Orange lines: significant relationship only in SZ group. **F**. Correlations between PL/V1 (left Y-axis) and PL/STSdp (right Y-axis). Schematic of proposed V1-➔PL-➔STSdp path. Beta coefficients for direct and indirect path (a*b) paths given. **G**. Correlations and TPOJ1-➔PM-➔STSdp mediated path. **H**. Correlations and PI-➔MST-➔TPOJ1 path.

Subcortically, PulN and V1 activation were significantly correlated across participants (F(1,54)=20.43, p<.001 Adj. R^2^=.364) (**Figure 3D**). In Sz patients, PulN activity also predicted MST (F(1,54)=6.40, p=.014; Adj. R^2^=.193), TPOJ1 (F(1,54)=8.51, p=.005; Adj. R^2^=.385) and STSdp (F(1,54)=9.03, p=.004; Adj. R^2^=.271) activation (**Figure 3E)**. Exploratory mediation analyses involving PulN subdivisions were used to further investigate these pulvino-cortical interrelations in Sz, based on observed pairwise correlations (**Supplementary Figure S3A)** and known anatomical projection patterns.

Consistent with “visual PulN” concepts, activation of V1 significantly predicted that of PL (Beta=.42; CI:[0.18, 0.66]), which in turn was a significant predictor of STSdp activation (Beta=1.06; CI:[0.18, 0.66]) (**Figure 3F**). After controlling for PL activation, V1 was no longer associated with STSdp (Beta=.10; CI:[-0.41, 0.60]). Moreover, using a bootstrapping approach, the (unstandardized) coefficient for the indirect pathway from V1-➔PL-➔STSdp was significant (Beta=.45; CI:[0.08, 0.84]), consistent with full mediation. Although PM also potentially mediated the path between V1 and STSdp (Beta=.32; CI:[0.01, 0.64]), the proportion of the effect of V1 on STSdp activation mediated through PM (total effect/indirect effect) was lower (58%) than the corresponding proportion through PL (82%) (**Supplementary Table 2)**.

A significant mediating effect of PM was also observed between TPOJ1 and STSdp (Beta=.59; CI:[0.01, 1.13]). Specifically, whereas the relationship between both TPOJ1 and PM (Beta=.47; CI:[0.21, 0.73]) and PM and STSdp (Beta=1.26; CI:[0.29, 2.22]) was significant (**Figure 3G**), after controlling for the proposed mediator, the TPOJ1-STSdp association was not significant (Beta=0.12; CI:[-0.64, 0.88]), again suggesting full mediation.

Lastly, activation of PI significantly predicted MST activity (Beta=0.42; CI:[0.12, 0.73]) which in turn predicted TPOJ1 (Beta=0.62; CI:[0.28, 0.95]) (**Figure 3H**). The analysis found no significant association between PI and TPOJ1 when conditioning on MST (Beta=0.13; CI:[-0.16, 0.41]), but did find a significant indirect pathway, consistent with full mediation of PI effects by MST (Beta=0.26; CI:[0.13, 0.60]).

### ASD

ASD participants showed similar impairments to Sz patients in higher-tier regions, including FFC (p=.020) and STS (TPOJ1 (p=.004); STSdp, p=.040) (**Figure 4A, B**). In STSdp the deficits, as in Sz, were greatest for dynamic emotional faces (3-way interaction: F(1,48)=5.73, p=.021) (**Figure 4C**) and predicted impaired FER (F(1,18)=7.39, p=.014; Adj. R^2^=.252) (**Figure 4D**). Additional activation deficits that were not observed in Sz were observed in more anterior parts of STS (STSva, p=.032) and frontal eye fields (FEF, p=.045) (**Supplementary Table 1**).

**Figure 4.**
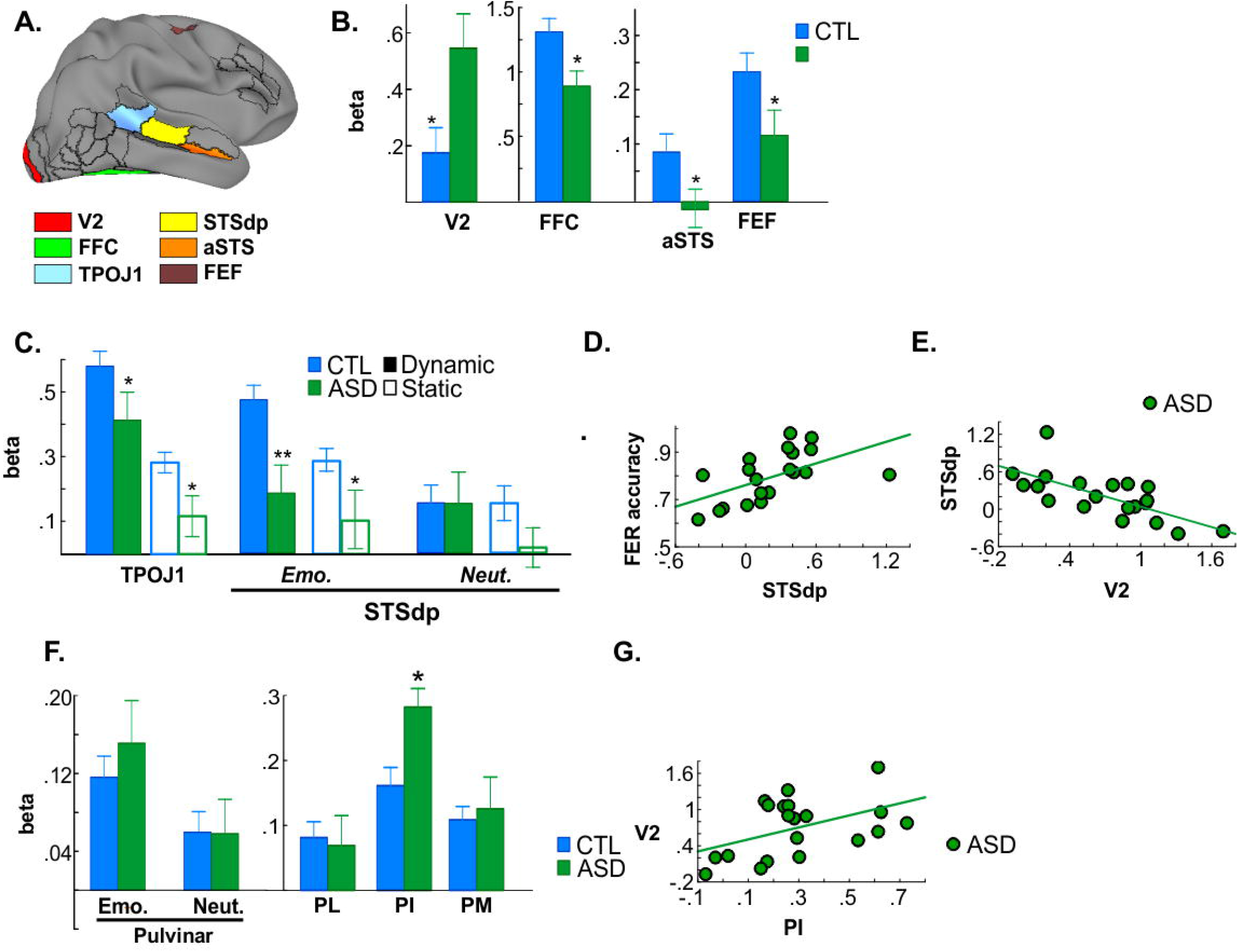
**A**. Localization of parcels with CTL versus ASD differences. **B**. Mean activation in CTL (blue) and ASD (green) groups. **C**. Mean activation to dynamic (solid)/static (open) faces in STS parcels. **D**. Correlation between STSdp deficits and FER in ASD. **E**. Correlation between enhanced V2 and reduced STSdp activation. **F**. Whole-pulvinar activation to emotional/neutral faces and mean activation within pulvinar subdivisions. **G**. Correlation between enhanced PI/V2 activation.

In addition, a differential pattern was observed within lower-tier areas in ASD versus Sz, including an overall increase in activity within early visual cortex (V2) in ASD that was significant relative to both control (F(1,48)=5.15, p=.028) and Sz (F(1,46)=4.57, p=.038) subjects. This heightened V2 activation specifically predicted impaired STSdp activity in ASD (F(1,18)=17.39, p=.001; Adj. R^2^=.463) (**Figure 4E**).

Also in contrast to Sz, activation of PulN did not differ overall between ASD and control participants (F(1,48)=.26, p=.614) (**Figure 4F**), nor did activation of PL or PM independently. By contrast, mean PI activation was significantly increased in ASD participants and was reflected in a significant group X subdivision interaction compared to neurotypical controls (F(2,47)=4.36, p=.011). Furthermore, increased activity of PI significantly predicted enhanced V2 activation in ASD (F(1,18)=7.14, p=.016; Adj. R^2^=.244) (**Figure 4G**). The increase in PI activation was again in contrast to that observed in Sz (F(1,46)=5.84, p=.020), reflecting a disorder-specific, double-dissociation of subnucleus-specific activation patterns in Sz versus ASD.

### Amygdala activation

Compared to control subjects, mean activation of the amygdala was significantly higher in Sz patients (F(1,56)=7.58, p=.008) and the group difference was larger for emotional, relative to neutral, faces (F(1,56)=4.93, p=.030). In the ASD group, activation of the amygdala was equivalent, overall, to that of control subjects (F(1,48)=1.39, p=.244) (**Supplementary Figure 2D**). Nevertheless, no correlations between amygdala activation and FER were observed either across of within groups (p>.09, all).

## Discussion

Deficits in social cognition contribute disproportionately to impaired functional outcome across a range of neurocognitive disorders, including Sz and ASD. FER is an important component of these deficits and, in the visual system, depends upon coordinated function of both subcortical and cortical regions for processing of static and dynamic facial features. Here, we investigated cortical and subcortical correlates of FER impairments in adults with Sz and ASD using a dynamic/static FER task that engages motion-sensitive areas as well as traditional face-processing regions. In addition, we investigated the subcortical pathway to cortex involving PulN.

The primary findings of the study relate to the relative involvement of cortico-cortical versus thalamo-cortical transmission paths underlying impaired FER in Sz. Traditionally, it was assumed that cortical regions showing intercorrelated activity mediate their joint activations primarily through direct cortico-cortical connections. More recent models by contrast propose that connections are mediated primarily by successive loops between cortex and thalamus, with higher-tier thalamic regions such as PulN and dorsomedial nuclei generally interacting with posterior and anterior association regions, respectively^49,50^. Within PulN, discrete subnuclei interact with specific visual cortical regions^29^. This theory converges with anatomical studies showing reduced PulN volume and cell number in schizophrenia^51-53^, along with our recent observations of impaired PulN activation to simple visual stimuli in Sz^14,15,54^.

In the present study, activation deficits in Sz were observed within the HCP-MMP1.0^45^ parcels comprising lower-tier visual regions including primary (V1), early visual (V4) and motion-sensitive cortex, along with higher-tier (multisensory) regions associated with FER. Within STS two discrete parcels were activated by the task– STSdp and TPOJ1. Activation of the STSdp, in particular, showed uniquely greater activation to dynamic emotional faces which correlated with behavioral measures of FER, in accord with the prominent role of STS in face-emotion assessment^55^. In Sz, STSdp deficits intercorrelated with impairments in activation of both early visual cortical regions and PulN.

### Role of pulvinar subdivisions

PulN is divided into discrete anatomical subdivisions which mirror the dorsal/ventral stream distinction of visual cortex^34^ such that the more lateral regions (PL) project predominantly to primary visual cortex and ventral visual stream, whereas a subset of nuclei in the inferior subdivision (PI) project mainly to dorsal stream regions including motion-sensitive cortex (e.g. MST)^28,34,56^. The medial subdivision (PM) is primarily connected with multimodal sensory association areas as well as prefrontal and cingulate cortices and has been tied to emotion processing^35^. In the present study, in addition to intercorrelated cortical activation deficits, we observed correlations between cortical regions and PulN subnuclei. We therefore conducted a series of mediation analyses to evaluate underlying pathways.

An initial analysis evaluated the potential pathways underlying intercorrelated deficits between V1 and STS in the schizophrenia group. A significant indirect pathway from V1-➔PL-➔STSdp was observed, in support of indirect mediation by PL. A possible indirect pathway from V1-➔PM-➔STSdp was also detected, but on the assumption that these two pathways operate independently, the pathway involving PM accounted for a smaller proportion of the effect of V1 on STSdp.

Consistent with mediation by PM, the intercorrelation between the STS parcels (TPOJ1 and STSdp) was not significant once an indirect path via PM was modeled. In contrast, PI appeared to mediate its effects via MST. Of note, unlike PL and PM, activation of PI was relatively intact in patients. Given that PI receives much of its driving inputs from neurons in the superior colliculus^34^, this finding is suggestive of unimpaired input via the retinotectal system. By contrast, the observed deficits in V1 are consistent with impaired input via the geniculostriate visual pathway^14,57^.

Although anatomical abnormalities in PulN are well documented in schizophrenia^53,58^, the functional consequences of these abnormalities have, to date, remained relatively unexplored. Here, we provide evidence that impairments in visual PulN function significantly undermines visual processing required for effective face processing. In specific, deficits in PL function may mediate effects of impaired V1 activation, which, in turn likely reflects impaired magnocellular input to cortex. In addition, impaired PM activation mediated impaired input from more posterior to mid-STS regions, suggesting that accumulating deficits across successive cortico-pulvinar loops may lead to the large effect-sized deficits in FER-related reduced STS activation in Sz.

### Comparison to ASD

While deficits in social cognition are a prominent component of Sz, they are not unique to the disorder. In particular, we^14^ and others^59,60^ have reported FER deficits in adult ASD subjects that are as severe as those observed in Sz, despite the much higher level of overall function. Consistent with these prior results, adult ASD participants in the present study showed FER and STS activation deficits that were similar to those of Sz, supporting a role for STS as a common mediator of FER dysfunction across disorders.

Despite the similar STS impairments, ASD participants showed markedly different patterns of disturbance within early visual regions, including normal V1 activation patterns but increased PI and V2 activation compared with both Sz participants and controls. Although the source of the increased activation in ASD is not known, a parsimonious explanation would be hyperactivity of the subcortical retinocollicular pathway, which provides preferential input to PI^34^ and which, in turn, acts like a driver to V2^33^ (see **Supplement** for further discussion). Regardless of underlying pathophysiology, the differential mechanisms suggest that neural mechanisms underlying FER deficits in Sz are disorder-specific, and, consistent with RDoC concepts, suggest the need to dissect pathophysiological mechanisms across units of analysis^61^.

### Amygdala activation

Lastly, in the present study amygdala activation was elevated overall in Sz, as reported previously (reviewed in^62^), possibly reflecting abnormal salience attribution to neutral stimuli^63-65^, heightened anxiety^66^ and/or paranoid ideation^67^.

## Limitations

Despite our differential findings, some limitations must be considered. First, Sz participants were receiving antipsychotic medication which may have impacted measures on brain activity. We did not observe any correlations with medication dose (see **Supplementary Material**), however, this issue could be best addressed in future studies involving medication-naïve patients. Additionally, we did not track fixation locations either in the scanner or during behavior. Thus, we do not know if activation failures relate to inability to process information, or simply from differential facial scanning approaches. Lastly, Sz participants had lower IQ and were older than controls or ASD participants, although correcting for IQ and age did not diminish the findings.

## Conclusions

In summary, higher cortical (e.g. STS) contributions to impaired FER have been extensively documented in Sz and ASD^1^, but early visual and subcortical contributions have been evaluated to only a limited degree. Here, we demonstrate significant but opposite abnormalities of circuits centered on PulN in Sz versus ASD that correlate with impaired STS activation, which in turn correlated across groups with impaired FER. These findings highlight the importance of close integration between subcortical and cortical visual processing pathways and the potential breakdown of this tight coordination in Sz and ASD. Further, the findings reaffirm that similar behavioral deficits (e.g. impairment in social cognition) do not necessarily imply convergent pathophysiological mechanisms, and that physiological measures may be useful for guiding etiological and interventional studies in neuropsychiatry.

## Data Availability

The data that support the findings of this study are available on request from the corresponding author (AM). The data are not publicly available due to their containing information that could compromise the privacy of research participants.

## Acknowledgements

The authors thank the staff of the Clinical Research and Evaluation Facility and the Clinical Evaluation Center at the Nathan S. Kline Institute for Psychiatric Research and all research participants for their contributions.

---

**Table 1:** Participant characteristics. Control (CTL), schizophrenia (SZ) autism spectrum disorder (ASD). Chlorpromazine equivalents (CPZ); *PSI:* Processing Speed Index; *POI:* Perceptual Organization Index; *ER-40*: Penn Emotion Recognition Task; *PANSS*: Positive and Negative Syndrome Scale; *ADOS-2*: Autism Diagnostic Observation Schedule, Second Edition, Communication (Comm.) and Social Interaction (Social Inter.) scores. Asterisks denote significant differences between CTL and SZ participants; plus signs denote differences between CTL and ASD participants (**/++p<.01; ***/+++p<.001). Standard deviations in parentheses.

|  | CTL | SZ | ASD |
| --- | --- | --- | --- |
| *Age* | 31.4 (9.8) | 38.8 (10.1)** | 28.9 (7.7) |
| *Gender (F/M)* | 8/22 | 8/20 | 4/16 |
| *Years of Education* | 14.6 (2.0) | 11.8 (2.0)*** | 13.7 (2.5) |
| *Participant SES* | 39.6 (11.5) | 24.4 (6.3) *** | 33.7 (11.8) |
| *Parental SES* | 46.1 (14.8) | 38.3 (13.1) | 49.2 (11.5) |
| *IQ* | 103.1 (8.9) | 96.7 (9.9)* | 101.2 (9.0) |
| *PSI* | 101.5 (12.3) | 81.8 (12.7)*** | -- |
| *POI* | 108.9 (16.1) | 88.7 (15.7)*** | -- |
| *Illness Duration (yrs)* | -- | 14.2 (9.1) | -- |
| *CPZ Equiv. SZ)* | -- | 834.8 (629.1) | -- |
| **Anti-Psychotic Medication Type** |  | Typical (6) |  |
|  |  | Atypical (15) |  |
|  |  | Combination (7) |  |
| *ER-40* | 35.8 (1.9) | 28.7 (5.5)*** | 30.2 (4.7)+++ |
| *PANSS (positive)* | -- | 11.1 (4.1) | -- |
| *PANSS (negative)* | -- | 19.3 (5.6) | -- |
| *ADOS-2 (Comm.)* | -- | -- | 4.7 (1.5) |
| *ADOS-2 (Social Inter.)* | -- | -- | 8.3 (2.5) |

