## Supplementary Material for "Disease-specific contribution of pulvinar dysfunction to impaired emotion recognition in schizophrenia"

**Supplementary Materials and Methods**

**Participants**

Seventy-eight participants took part, including 28 participants diagnosed with schizophrenia (Sz) using the Structured Clinical Interview for DSM-IV[1], 20 adults with autism spectrum disorder (ASD), confirmed by the Autism Diagnostic Observation Schedule, Second Edition, and 30 neurotypical controls. All Sz participants were on a stable dose of antipsychotic medication. All participants had at least 20/22 corrected visual acuity on a Logarithmic Visual Acuity Chart. On average, Sz participants were older ($F(1,56)=7.24$, $p=.009$) and had lower IQ scores ($F(1,56)=6.54$, $p=.013$) than controls. Participants were recruited from the central research database and volunteer recruitment pool at the Nathan Kline Institute for Psychiatric Research (NKI). All ASD participants and a subset of 19 Sz and 17 controls participated in our previous EEG/fMRI study of visual sensory dysfunction as reported in[2], which did not include data from the present paradigm.

**Behavioral FER Measures**

A forced-choice behavioral task was administered following the fMRI scan using both static and dynamic emotional faces (80 stimuli total; neutral faces were not included). After each presentation, subjects were prompted to press one of five buttons to indicate if the actor's expression was 1) happy, 2) sad, 3) angry, 4) fearful or 5) none of the above. Accuracy, as opposed to response time was emphasized. The trial ended when subjects responded. The Penn Emotion Recognition (ER-40) test[3] was also administered to participants and its results compared to those of the present FER paradigm. ER-40 was not available from one control participant.

**Imaging Acquisition**

All imaging took place on a Siemens 3T TiM Trio scanner housed at NKI's Center for Advanced Brain Imaging. On each functional scan, two-hundred-twenty T2*-weighted echo-planar images were acquired in the axial plane (TR=2000ms;TE=38ms;FA=90°; voxel size = 27.0 $mm^3$; 32 slices). At least one high-resolution structural image of the entire brain was acquired from each participant using an MPRAGE sequence (TR=2500 ms, TE=3.5 ms, TI=1200 ms, matrix=256x256, voxel size=1.0$mm^3$, 192 slices).

Individual cortical surfaces were rendered with Freesurfer (http://surfer.nmr.mgh.harvard.edu/) and registered to the std.141 fsaverage mesh with SUMA (https://afni.nimh.nih.gov/Suma). Segmentation of the thalamic nuclei (to derive pulvinar) [4]and amygdala[5] was carried out using automatic segmentation tools incorporated in Freesurfer.

**Imaging Analyses**

Data were preprocessed using the AFNI afni_proc.py function consisting of concatenating data from two runs, removal of signal deviation >2.5 SDs from the mean (AFNI's 3dDespike), temporal alignment, identification of motion outliers per run, spatial smoothing with a 6mm full width at half maximum Gaussian kernel and scaling of blood-oxygen-level-dependent (BOLD) values to mean percent signal change[6]. Single-participant statistical analyses were conducted within the framework of the general linear model (GLM). The GLM model included regressors for each stimulus type (emotional dynamic, emotional static, neutral dynamic, neutral static) as well as regressors for the six motion parameters (three rotations, three translations) and their first derivatives, per run. Time points with large head motion between successive time points were censored. Surface-based analyses were carried out on the gray-matter ordinates of each individual cortical surface aligned to the Freesurfer 141-standard mesh. To assess activation of pulvinar and amygdala, identical analyses were carried out in the individual native-space volumes.

Cortical data was sampled to the Human Connectome Project multimodal cortical parcellation (HCP-MMP1.0)[7] which delineates 180 brain parcels per hemisphere based on functional and structural properties (**Supplementary Figure 1A**). Functional activations were analyzed within a 35-parcel mask (**Supplementary Figure 1B**) consisting of parcels with significant activation ($p<.001$, uncorrected) across all subjects and stimuli. To assess activation of subcortical structures (pulvinar and amygdala), identical statistical analyses were carried out in the individual native-space volumes.

**Clinical correlations**

No significant correlations were observed between behavioral performance or cortical/subcortical activation patterns and medication dose (CPZ equivalents) in Sz patients. Functional activation strengths did not correlate with measures of general cognitive ability (PSI and IQ) in any group ($p>.11$ for all), however, in Sz ($r=.378$, p=.049) and control (r=.466, p=.044) participants, perceptual organization skill (POI) correlated with performance on the FER task as well as with STSdp (HC: r=.539, p=.017; Sz: r=.399, p=.035).

**Discussion**

*Sz vs ASD:* Despite the convergent deficits in the STS region, a significantly divergent pattern of abnormality was observed in earlier tiers of the visual system. In the case of Sz, significant impairment in activation of striate (V1) visual cortex was observed, which correlated with impaired STSdp activation. By contrast, in ASD, markedly increased responses within the early visual system (V2) and an opposite slope of the relationship between V2 and STSdp activation were observed. Activation within other task-activated visual regions, including V1 and MST, was unaffected in ASD, echoing our recent study in which response amplitudes were also normal within V1, but increased in early visual and dorsal visual regions[2]. Similar visual hypo/hyper activation patterns in Sz versus ASD have been observed in both fMRI (reviewed in [8, 9]) and electrophysiological[2, 10-12] studies, supporting the concept that dysregulation of the early visual system may undermine later stages of visual processing.

Patterns of subcortical activation also distinguished between ASD and Sz participants. In particular, wheras PulN activity was markedly reduced in Sz, activation of the inferior PulN subdivision was significantly elevated in ASD participants, in line with findings from our previous studies[2, 11] and those of others[2, 13, 14]. Although the source of the increased activation is not known, a parsimonious explanation would be hyperactivity of the subcortical retino-collicular pathway, which provides preferential input to PI[15] and which, in turn, acts like a driver to V2[16]. In humans, this system typically weakens with age as the retinogeniculate system increases in functionality (reviewed in[17]). Abnormal persistence of this system into adulthood could thus underlie the activation disturbance pattern observed in ASD. In ASD, unlike schizophrenia, we found no evidence for impairments in function of other pulvinar subdivisions. By contrast, in Sz we observed normal activation patterns in PI but not other PulN regions, suggesting relative intact input via the retinotectal system.

Overall, these findings support the concept that dysregulation of the early visual system, whether in the direction of increased or decreased activation, may undermine later stages of visual processing and further highlight the importance of sensory processing abnormalities to the pathophysiology of social cognitive impairment across neuropsychiatric disorders.

**Supplementary Tables and Figure Legends**

| | HC | SZ | ASD | HC v SZ | | HC v ASD | | ASD v SZ | |
|---|---|---|---|---|---|---|---|---|---|
| | | | | F(1,56) | p | F(1,48) | p | t(46) | p |
| **V1** | 0.56(0.32) | 0.30(0.31) | 0.37(0.45) | 10.41 | .002* | 3.16 | 0.082 | 0.67 | 0.506 |
| **V2** | 0.06(0.50) | 0.06(0.72) | 0.45(0.61) | 0.01 | 0.989 | 5.15 | .028* | 1.79 | 0.039* |
| **V3** | 0.36(0.27) | 0.31(0.30) | 0.42(0.34) | 0.50 | 0.484 | 0.38 | 0.541 | 1.14 | 0.260 |
| **V4** | 1.47(0.59) | 1.07(0.40) | 1.45(0.81) | 8.63 | .005* | 0.01 | 0.921 | 2.10 | .041* |
| **V8** | 0.63(0.40) | 0.66(0.40) | 0.83(0.64) | 0.07 | 0.788 | 1.93 | 0.171 | 1.17 | 0.247 |
| **FFC** | 1.32(0.56) | .99(0.52) | .89(0.53) | 4.20 | .045* | 5.83 | 0.020* | 0.65 | 0.520 |
| **PIT** | 1.35(0.51) | 1.31(0.63) | 1.34(0.85) | 0.06 | 0.811 | 0.00 | 0.977 | 0.14 | 0.886 |
| **VVC** | 0.40(0.29) | 0.51(0.32) | 0.44(0.46) | 1.62 | 0.208 | 0.10 | 0.758 | -0.62 | 0.535 |
| **MST** | 0.46(0.20) | 0.35(0.21) | 0.48(0.26) | 4.30 | .042* | 0.20 | 0.676 | 2.09 | .042* |
| **LO2** | 1.03(0.51) | 1.11(0.44) | 1.03(0.68) | 0.47 | 0.494 | 0.00 | 0.976 | -0.51 | 0.614 |
| **MT** | 0.37(0.22) | 0.46(0.37) | 0.51(0.28) | 1.50 | 0.222 | 3.80 | 0.058 | 0.42 | 0.679 |
| **PH** | 0.40(0.29) | 0.49(0.38) | 0.40(0.34) | 0.96 | 0.332 | 0.00 | 0.998 | -0.81 | 0.422 |
| **V4t** | 0.75(0.41) | 0.94(0.44) | 0.89(0.55) | 3.07 | 0.085 | 1.12 | 0.295 | -0.35 | 0.727 |
| **FST** | 0.00(0.15) | -0.03(0.18) | 0.00(0.21) | 0.44 | 0.508 | 0.00 | 0.983 | 0.54 | 0.593 |
| **FEF** | 0.24(0.17) | 0.28(0.24) | 0.12(0.21) | 2.14 | 0.149 | 4.24 | 0.044* | -2.50 | .016* |
| **STSda** | 0.10(0.15) | 0.12(0.20) | 0.06(0.19) | 0.17 | 0.685 | 0.91 | 0.344 | -1.12 | 0.270 |
| **STSdp** | 0.19(0.15) | 0.07(0.22) | 0.08(0.23) | 5.81 | .019* | 4.48 | .040* | 0.10 | 0.923 |
| **STSvp** | 0.04(0.15) | -0.04(0.31) | -0.01(0.19) | 1.55 | 0.219 | 1.08 | 0.304 | 0.36 | 0.724 |
| **STSva** | 0.09(0.17) | 0.15(0.34) | -0.01(0.16) | 0.76 | 0.388 | 4.88 | 0.032* | -2.03 | .048* |
| **FOP5** | 0.10(0.14) | 0.09(0.22) | 0.19(0.20) | 0.02 | 0.901 | 3.45 | 0.069 | 1.54 | 0.131 |
| **TE2p** | 0.25(0.31) | 0.31(0.43) | 0.22(0.42) | 0.39 | 0.535 | 0.08 | 0.774 | -0.72 | 0.473 |
| **PHT** | 0.15(0.22) | 0.17(0.29) | 0.09(0.23) | 0.07 | 0.786 | 0.91 | 0.346 | -1.03 | 0.308 |
| **STV** | 0.27(0.16) | 0.17(0.21) | 0.22(0.27) | 4.56 | .037* | 0.85 | 0.360 | 0.69 | 0.494 |
| **TPOJ1** | 0.51(0.24) | 0.28(0.27) | 0.27(0.33) | 11.83 | .001* | 9.43 | .004* | -0.19 | 0.852 |
| **TPOJ2** | 0.35(0.25) | 0.34(0.28) | 0.28(0.27) | 0.00 | 0.958 | 0.69 | 0.410 | -0.72 | 0.476 |
| **TPOJ3** | 0.15(0.20) | 0.21(0.26) | 0.18(0.18) | 0.85 | 0.360 | 0.19 | 0.664 | -0.47 | 0.638 |
| **LIPd** | 0.25(0.21) | 0.27(0.24) | 0.24(0.29) | 0.04 | 0.845 | 0.03 | 0.872 | -0.30 | 0.767 |
| **IP1** | 0.19(0.24) | 0.13(0.24) | 0.18(0.26) | 1.16 | 0.287 | 0.03 | 0.856 | 0.75 | 0.457 |
| **IP0** | 0.08(0.14) | 0.19(0.28) | 0.07(0.29) | 3.21 | 0.079 | 0.02 | 0.894 | -1.34 | 0.186 |
| **45** | 0.14(0.14) | 0.09(0.19) | 0.11(0.27) | 1.08 | 0.303 | 0.27 | 0.607 | 0.23 | 0.816 |
| **IFJa** | 0.42(0.25) | 0.43(0.33) | 0.39(0.37) | 0.01 | 0.918 | 0.12 | 0.736 | -0.37 | 0.713 |
| **IFJp** | 0.36(0.21) | 0.40(0.33) | 0.35(0.29) | 0.24 | 0.626 | 0.05 | 0.830 | -0.55 | 0.585 |
| **IFSp** | 0.34(0.19) | 0.23(0.24) | 0.26(0.29) | 4.05 | .049* | 1.42 | 0.240 | 0.41 | 0.685 |
| **IFSa** | 0.17(0.15) | 0.14(0.23) | 0.13(0.23) | 0.24 | 0.630 | 0.56 | 0.458 | -0.23 | 0.819 |
| **p9-46v** | 0.17(0.14) | 0.15(0.23) | 0.13(0.18) | 0.10 | 0.755 | 0.84 | 0.363 | -0.44 | 0.662 |

**Supplementary Table 1:** Mean beta parameter values in each of the 35 parcels shown in **Supplementary Figure 1C** for the control (CTL), schizophrenia (SZ) and autism (ASD) groups. Standard errors of the mean are in parentheses. F- and p-values for the main effect of group membership in the ANOVAs contrasting CTL vs SZ, CTL vs ASD and ASD vs SZ. Asterisks indicate statistical significance.

| | | Path | B | SE | t | p | 95% CI | |
|---|---|---|---|---|---|---|---|---|
| | | | | | | | LL | UL |
| | **V1-PL** | a | 0.42 | 0.12 | 3.65 | 0.001 | 0.18 | 0.66 |
| ***X=V1*** | **PL-pSTS** | b | 1.06 | 0.34 | 3.13 | 0.005 | 0.36 | 1.76 |
| ***Y=pSTS*** | **V1-pSTS** | c | 0.54 | 0.23 | 2.36 | 0.026 | 0.07 | 1.02 |
| ***M=PL*** | **V1-pSTS l PL** | c' | 0.10 | 0.25 | 0.40 | 0.694 | -0.41 | 0.60 |
| | **Indirect** | a*b | 0.45 | 0.20 | -- | -- | 0.08 | 0.84 |
| | **V1-PM** | a | 0.28 | 0.10 | 2.90 | 0.007 | 0.08 | 0.47 |
| ***X=V1*** | **PM-pSTS** | b | 1.15 | 0.43 | 2.69 | 0.013 | 0.27 | 2.03 |
| ***Y=pSTS*** | **V1-pSTS** | c | 0.54 | 0.23 | 2.36 | 0.026 | 0.07 | 1.02 |
| ***M=PM*** | **V1-pSTS l PM** | c' | 0.23 | 0.24 | 0.95 | 0.351 | -0.26 | 0.72 |
| | **Indirect** | a*b | 0.32 | 0.16 | -- | -- | 0.01 | 0.64 |
| | **TPJ-PM** | a | 0.47 | 0.12 | 3.75 | 0.001 | 0.21 | 0.72 |
| ***X=TPJ*** | **PM-pSTS** | b | 1.26 | 0.47 | 2.69 | 0.013 | 0.29 | 2.22 |
| ***Y=pSTS*** | **TPJ-pSTS** | c | 0.71 | 0.33 | 2.14 | 0.042 | 0.03 | 1.39 |
| ***M=PM*** | **TPJ-pSTS l PM** | c' | 0.12 | 0.37 | 0.32 | 0.748 | -0.64 | 0.88 |
| | **Indirect** | a*b | 0.59 | 0.29 | -- | -- | 0.00 | 1.13 |
| | **PI-MST** | a | 0.42 | 0.15 | 2.88 | 0.008 | 0.12 | 0.73 |
| ***X=PI*** | **MST-TPJ** | b | 0.62 | 0.16 | 3.81 | 0.001 | 0.28 | 0.95 |
| ***Y=TPJ*** | **PI-TPJ** | c | 0.39 | 0.15 | 2.60 | 0.015 | 0.08 | 0.69 |
| ***M=MST*** | **PI-TPJ l MST** | c' | 0.13 | 0.14 | 0.92 | 0.369 | -0.16 | 0.41 |
| | **Indirect** | a*b | 0.26 | 0.12 | -- | -- | 0.13 | 0.60 |

**Supplementary Table 2:** Results of mediation analyses testing whether a proposed causal effect of X (predictor) on Y (outcome) may be transmitted through a mediating (M) variable. For each mediation analysis, the coefficients (B), standard error (SE), t-statistic, p-value and lower (LL) and upper (UL) levels for the 95% confidence interval (CI) are given for the direct paths: (a) X and M; (b) M and Y; (c) X and Y; (c') X and Y, conditional on M and for the indirect (a*b) effect.

**Supplementary Figure 1**: **A.** The HCP-MMP1.0 parcellation atlas[7] projected on the semi-inflated fsaverage (std.141) template brain. Borders of 180 parcels per hemisphere in black. **B.** Whole-brain beta parameter maps of activation elicited by all stimuli and across all participants, superimposed on the template brain with borders of HCP parcels demarcated. **C.** Thirty-five parcels with significant activation across all subjects and all face stimuli, identified by a one-sample t-test vs. 0 (collapsed over all stimuli) covaried by participant age and thresholded at an (uncorrected) p-value of .001

**Supplementary Figure 2:** **A.** FER accuracy as a function of face-emotion type and representative face-emotion and neutral stimuli. Accuracy was equivalent in all three groups for happy faces. Fearful faces elicited the largest group difference in both SZ (orange; ($F(1,56)=8.89$, $p=.005$) and ASD (green; $F(1,48)=9.59$, $p=.004$) participants compared to the CTL (blue) group. **B.** Scores on the Penn Emotion Recognition (ER-40) test were lower in both SZ ($F(1,55)=29.60$, $p<.001$) and ASD ($F(1,47)=23.26$, $p<.001$) participants compared to the CTL group. **C.** Across participants, ER-40 scores significantly predicted mean accuracy on the FER task results ($F(1,72)=8.01$, $p=.006$; $R^2=.401$).

**Supplementary Figure 3:** **A.** Pairwise cross-correlation matrix of activation in cortical parcels with significant group differences and subcortical areas (pulvinar, PulN; amygdala, Amyg). CTL group is left side matrix, correlation matrix for SZ participants is on right. Significant within-group correlations, corrected for multiple comparisons, are indicated with white (CTL) and black (SZ) asterisks. **C.** Cross-correlation matrix for SZ group between activation of each pulvinar subdivision (lateral, PL; inferior, PI; medial, PM; anterior, PA) and cortical parcels and amygdala. We investigated whether the association between V1 and pSTS was mediated by PL (or PM), whether PM also mediated the association between TPJ and pSTS, and whether MST mediated the association between PI and TPJ. **B.** Localization of the amygdala on MNI template brain and bar plots of mean amygdala activation (beta parameter) for emotional and neutral faces in the CTL, SZ and ASD group. (*p<.05; **p<.01;***p<.005)

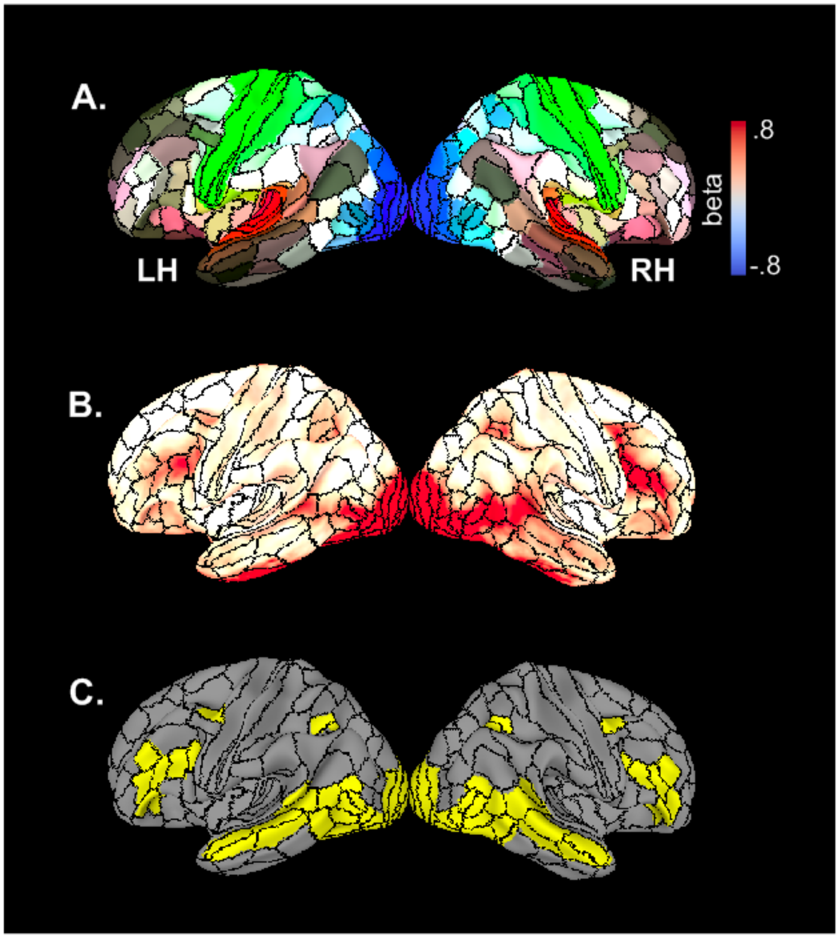


**Fig S1**

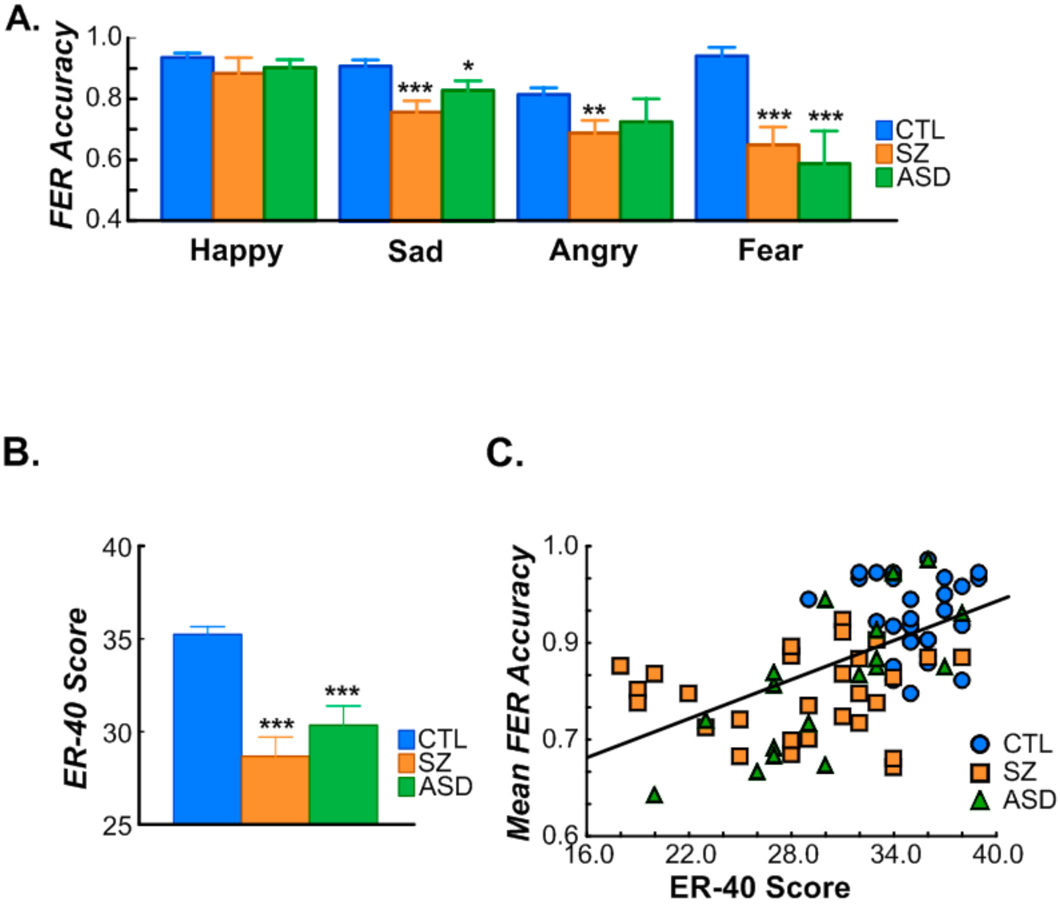
A.
FER Accuracy
1.0
0.8
0.6
0.4
***
*
**
***
***
CTL
SZ
ASD
Happy
Sad
Angry
Fear
B.
C.
40
35
30
25
ER-40 Score
***
***
CTL
SZ
ASD
Mean FER Accuracy
1.0
0.9
0.7
0.6
16.0
22.0
28.0
34.0
40.0
ER-40 Score
CTL
SZ
ASD

**Fig S2**

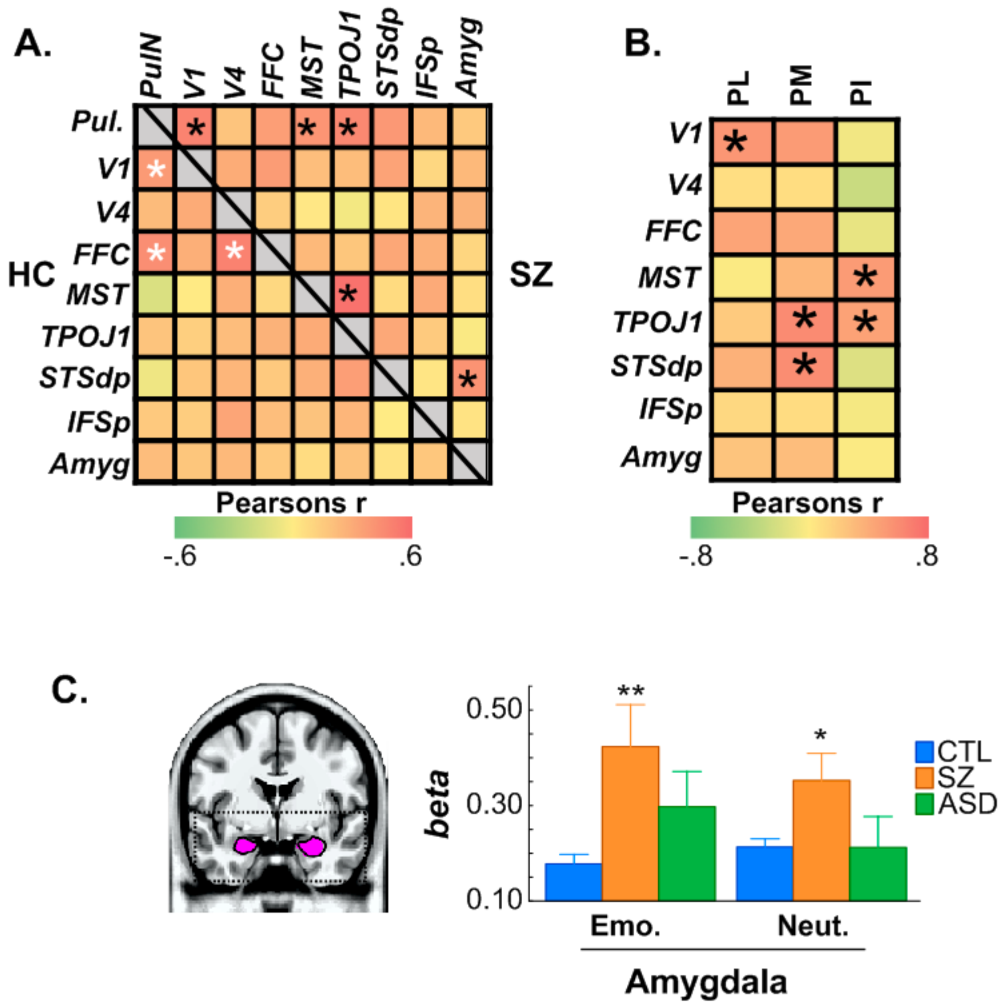
A.
PulN
V1
V4
FFC
MST
TPOJ1
STSdp
IFSp
Amyg
Pul.
HC
SZ
Pearsons r
-.6
.6
B.
PL
PM
PI
Pearsons r
-.8
.8
C.
0.50
0.30
0.10
beta
**
*
CTL
SZ
ASD
Emo.
Neut.
Amygdala

**Fig S3**
